# Comprehension of acoustically degraded speech in Alzheimer’s disease and primary progressive aphasia

**DOI:** 10.1101/2022.12.05.22283108

**Authors:** Jessica Jiang, Jeremy CS Johnson, Maï-Carmen Requena-Komuro, Elia Benhamou, Harri Sivasathiaseelan, Anthipa Chokesuwattanaskul, Annabel Nelson, Ross Nortley, Rimona S Weil, Anna Volkmer, Charles R Marshall, Doris–Eva Bamiou, Jason D Warren, Chris JD Hardy

## Abstract

Successful communication in daily life frequently depends on accurate decoding of speech signals that are acoustically degraded by challenging listening conditions. This process presents the brain with a demanding computational task that is vulnerable to neurodegenerative pathologies. However, despite recent intense interest in the link between hearing impairment and dementia, daily hearing measures (such as degraded speech comprehension) in these diseases remain poorly defined. Here we addressed this issue in a cohort of 19 patients with typical Alzheimer’s disease (AD) and 31 patients representing canonical syndromes of primary progressive aphasia (PPA), in relation to 25 healthy age-matched controls. As a model paradigm for the acoustically degraded speech signals of daily life, we used noise-vocoding: synthetic division of the speech signal into a variable number of frequency channels constituted from amplitude-modulated white noise, such that fewer channels convey less spectrotemporal detail thereby reducing intelligibility. We investigated the impact of noise-vocoding on recognition of spoken three-digit numbers and used psychometric modelling to ascertain the threshold number of noise-vocoding channels required for 50% intelligibility by each participant. Associations of noise-vocoded speech intelligibility threshold with general demographic, clinical and neuropsychological characteristics and regional grey matter volume (defined by voxel-based morphometry of patients’ brain MR images) were also assessed. Compared with healthy older controls, all patient groups had a significantly higher mean noise-vocoded speech intelligibility threshold, particularly marked in logopenic variant and nonfluent-agrammatic variant PPA and significantly higher in AD than in semantic variant PPA (all p<0.05). Noise-vocoded intelligibility threshold discriminated dementia syndromes (in particular, Alzheimer’s disease) well from healthy controls. Further, this central hearing measure correlated with overall disease severity but not with measures of peripheral hearing or clear speech perception. Neuroanatomically, after correcting for multiple voxel-wise comparisons in pre-defined regions of interest, impaired noise-vocoded speech comprehension across dementia syndromes was significantly associated (p<0.05) with atrophy of left planum temporale, angular gyrus and anterior cingulate gyrus: a cortical network widely implicated in processing degraded speech signals. Taken together, our findings suggest that the comprehension of acoustically altered speech captures a central process relevant to daily hearing and communication in major dementia syndromes, with novel diagnostic and therapeutic implications.

## INTRODUCTION

Successful communication in the world at large depends on our ability to understand spoken messages under non-ideal listening conditions. In our daily lives, we are required to interpret speech that is acoustically degraded by a wide variety of different ways – we regularly conduct conversations over background noise, adapt to suboptimal telephone and video connections and interpret unfamiliar accents. The processing of such degraded speech signals presents the brain with a challenging computational problem, whereby acoustic signals (or ‘auditory objects’) of interest must be disambiguated from interfering (and changing) noise.^1-3^ Because speech signals are critical for communication, decoding of degraded speech is generally the most functionally relevant index of hearing ability in daily life. This process, normally automatic and relatively effortless, is impaired in neurodegenerative disorders such as Alzheimer’s disease (AD) and the ‘language-led’ dementia syndromes of the primary progressive aphasia (PPA) spectrum.^4-8^

Hearing impairment has recently been identified as a major risk factor for dementia and a driver of cognitive decline and disability.^4,9,10^ While most studies addressing this linkage have focused on peripheral hearing function measured using the detection of pure tones,^4,11,12^ mounting evidence suggests that measures of central hearing (auditory brain) function and in particular, the comprehension of degraded speech signals, may be more pertinent.^6,8^ Large cohort studies have identified impaired comprehension of degraded messages as a harbinger of dementia.^7,13,14^ More specifically, AD has been shown to impact speech-in-noise perception ^15^ and identification of dichotic digits.^6,16-18^ This is likely to reflect, at least in part, a generic impairment of auditory scene analysis in AD, affecting the parsing of nonverbal as well as verbal information and linked to degeneration of the core temporo-parietal ‘default mode’ network targeted by AD pathology.^15,19-22^

Further, both AD and PPA syndromes impair comprehension of non-native accents,^23-26^ sinewave speech ^27,28^ and noise-interrupted speech,^29^ suggesting that neurodegenerative pathologies impair the processing of degraded speech signals more generally. However, the neural mechanisms responsible, the types of speech degradation that are implicated in everyday listening and the effects of different neurodegenerative pathologies have not yet been fully clarified. There are several grounds on which the processing of degraded speech may be especially vulnerable to neurodegenerative pathologies.^5^ Neuroanatomically, the processing of degraded speech signals engages distributed neural networks in peri-Sylvian, pre-frontal and posterior temporo-parietal cortices: these same brain networks are targeted preferentially in PPA, particularly the nonfluent/agrammatic variant (nfvPPA) and logopenic variant (lvPPA) syndromes.^5,27,30,31^ Computationally, the comprehension of degraded speech signals depends on precise yet dynamic integration of information across neural circuitry ^4,5,8,32,33^ and neurodegenerative pathologies are likely to blight these computations early and profoundly.

One widely used technique for altering speech signals experimentally is noise-vocoding, whereby a speech signal is divided digitally into discrete frequency bands (‘channels’), each filled with white noise and modulated by the amplitude envelope of the original signal.^34^ This procedure degrades the spectral content of the speech signal while preserving its overall longer range temporal structure. Using noise-vocoding, the level of intelligibility of the speech signal can be controlled: fewer channels is equivalent to less spectral detail available, leading to less intelligible speech. Among various alternative methods,^5^ noise-vocoding has certain attributes that make it attractive as a paradigm to study the effects of disease on the processing of degraded speech. Noise-vocoding has been widely studied and its behavioural and neuroanatomical correlates in the healthy brain are fairly well established.^34-40^ As an exemplar of acoustic degradation based on reduction of spectral information, it is broadly applicable to a variety of daily listening scenarios requiring decoding of ‘noisy’ speech signals (for example, a poor telephone or video-conferencing line). In contrast to speech-in-noise perception, comprehension of noise-vocoded speech depends intrinsically on auditory object (phonemic) decoding rather than selective attention. Further, noise-vocoding offers the substantial advantage of generating a quantifiable threshold for intelligibility of the degraded speech signal, based on the number of vocoding channels. This potentially allows for a more sensitive, graded and robust determination of deficit, enabling comparisons between diseases, tracking of disease evolution and assessing the impact of therapeutic interventions.

Noise-vocoding has been previously applied in a joint behavioural and magnetoencephalographic study of nfvPPA, to assess the brain mechanisms that mediate comprehension of degraded speech in the context of relatively focal cerebral atrophy.^41^ This work showed that patients with nfvPPA rely more on cross-modal cues to disambiguate vocoded speech signals, and have inflexible predictive decoding mechanisms, instantiated in left inferior frontal cortex. However, noise-vocoding has not been exploited as a tool to compare degraded speech perception in different neurodegenerative syndromes. More generally, the cognitive and neuroanatomical mechanisms that mediate the processing of degraded speech and their clinical resonance in this disease spectrum remain poorly defined.

Here, using noise-vocoding, we evaluated the comprehension of acoustically degraded spoken messages in cohorts of patients with typical AD and with all major syndromes of PPA, referenced to healthy older listeners. We assessed how the understanding of noise-vocoded speech was related to other demographic and disease characteristics. We further assessed the structural neuroanatomical associations of the noise-vocoded speech intelligibility threshold in AD and PPA, using voxel-based morphometry on patients’ brain MR images. Based on available evidence with noise-vocoded and other degraded speech stimuli in these disease populations,^5,30,31,41^ we hypothesised that both AD and PPA patients would have elevated thresholds for comprehending vocoded speech compared with healthy controls, and that this deficit would be more severe in nfvPPA and lvPPA than in other neurodegenerative syndromes. We further hypothesised that elevated intelligibility threshold in the patient cohort would be correlated with regional grey matter atrophy in left posterior superior temporal, inferior parietal and inferior frontal cortices previously implicated in the processing of noise-vocoded speech in the healthy brain.^34-40^

## MATERIALS AND METHODS

### Participants

Nineteen patients with typical amnestic AD, nine patients with lvPPA, 10 patients with nfvPPA and 12 patients with semantic variant primary progressive aphasia (svPPA) were recruited via a specialist cognitive clinic. All patients fulfilled consensus clinical diagnostic criteria with compatible brain MRI profiles and had clinically mild-to-moderate disease.^42,43^ No patients with pathogenic mutations were included. Twenty-five healthy older control participants with no history of neurological or psychiatric disorders were recruited from the Dementia Research Centre volunteer database. All participants had a comprehensive general neuropsychological assessment (Table 1). None had a history of otological disease, other than presbycusis; participants assessed in person at the research centre had pure tone audiometry, following a previously described procedure (details in Supplementary Material online).

**Table 1.**
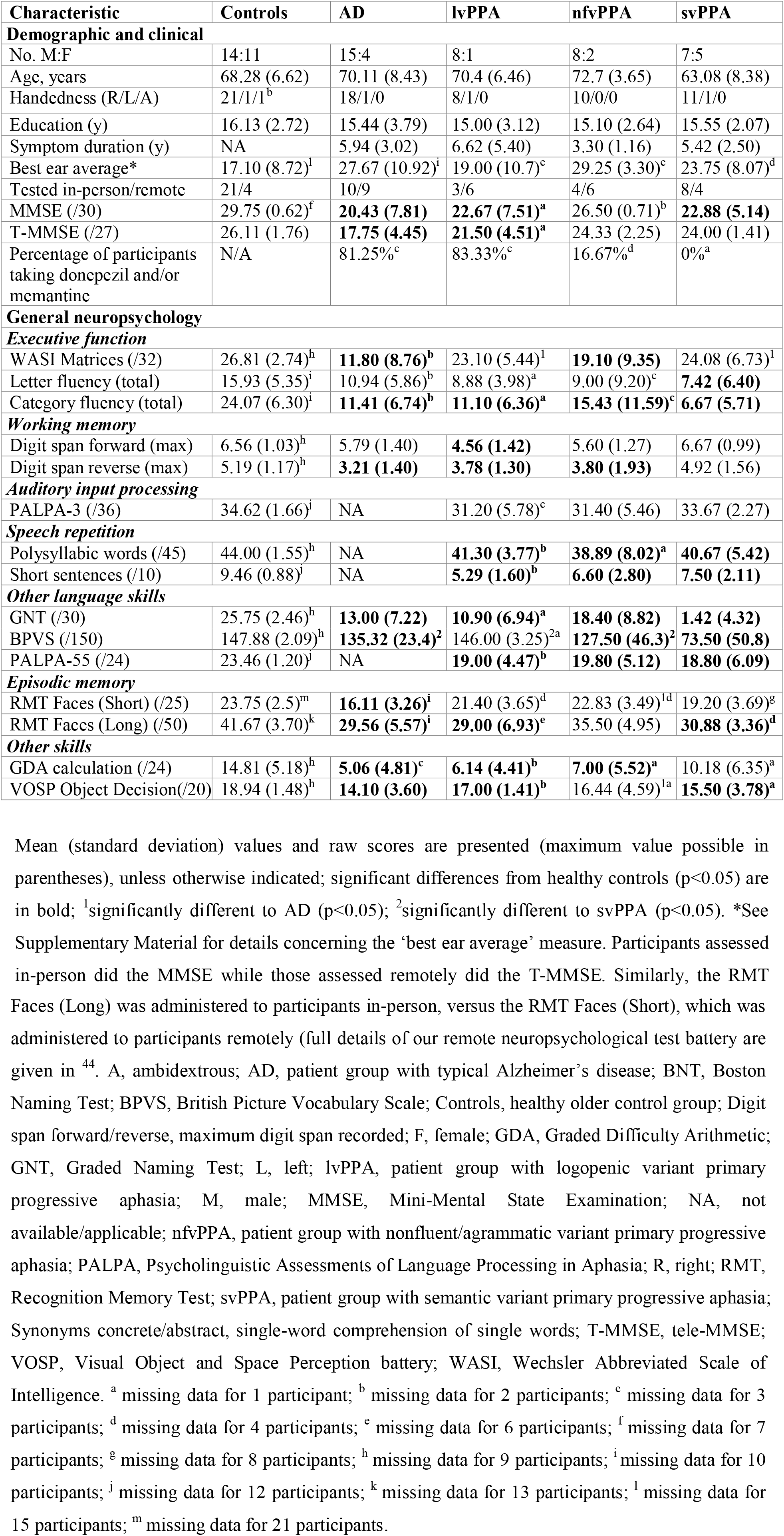
General demographic, clinical and neuropsychological characteristics of all participant groups

Due to the Covid-19 pandemic, some data for this study were collected remotely (see Supplementary Materials). We have described the design and implementation of our remote neuropsychological assessment protocol elsewhere.^44^

All participants gave informed consent to take part in the study. Ethical approval was granted by the UCL-NHNN Joint Research Ethics Committees, in accordance with Declaration of Helsinki guidelines.

### Creation of experimental stimuli

Lists of 50 different three-digit numbers (of the form, ‘five hundred and eighty seven’; examples in Supplementary Material online) were recorded by two young adult female speakers in a Standard Southern British English accent with neutral prosody. They were recorded in Audacity (v 2.2.3), using a condenser microphone with a pop-shield in a sound-proof booth. Speech recordings were noise-vocoded using Matlab® (vR2019b) (https://uk.mathworks.com/) to generate acoustically altered stimuli with a prescribed level of degraded intelligibility (see Figure S1 for spectrograms). Details concerning the synthesis of noise-vocoded stimuli are provided in Supplementary Material online. The vocoding intelligibility threshold for younger normal listeners is typically around three to four ‘channels’ ^34^; in this experiment, we noise-vocoded the speech recordings with one to 24 channels, sampling at each integer number of channels within this range to ensure we would be able to accurately capture even markedly abnormal psychometric functions in the patient cohort.

The final stimulus list comprised 100 different spoken three-digit numbers: four unvocoded (clear speech) and 96 noise-vocoded with four stimuli for each number of channels, ranging from one to 24.

### Experimental procedure

The stimuli were administered binaurally in a quiet room via Audio-Technica ATH-M50x headphones at a comfortable fixed listening level (at least 70 dB). Data for 30 participants were collected remotely via video link during the Covid-19 pandemic (see Table 1, further details in Supplementary Material online).

To familiarise them with the experimental procedure, participants were first asked to repeat five three-digit numbers (not included in the experimental session) that were spoken by the experimenter. Prior to presenting the experimental stimuli, participants were advised that the numbers they heard would vary in how difficult to understand they were, but that they should guess the number even if uncertain. Stimuli were presented in order of progressively decreasing channel number (intelligibility), from clear speech, then 24 to one vocoding channel. On each experimental trial, the task was to repeat the number (or as many of the three digits as the participant could identify). Participants were allowed to write down the numbers they heard rather than speaking them if preferred; in scoring, we accepted the intended target digit as correct, even if imperfectly articulated. Responses were recorded for offline analysis. During the experiment, no feedback about performance was given and no time limits were imposed.

### Analysis of clinical and behavioural data

Data were analysed in Matlab® (vR2019b) and in R® (v4). For continuous demographic and neuropsychological data, participant groups were compared using ANOVA and Kruskal Wallis tests (dependent on normality of the data); group categorical data were compared using Fisher’s exact tests. Performance profiles in seven healthy control participants who performed the experiment both in person and subsequently remotely were very similar, justifying combining participants tested in person and remotely in the main analysis (see Supplementary Material online). An alpha of 0.05 was adopted as the threshold for statistical significance on all tests.

Identification of noise-vocoded spoken numbers was scored according to the number of digits correct for each three-digit number (e.g., if the target number was ‘587’ and the participant responded ‘585’, they would score two points on that trial). As three digits were presented on every trial, this scoring effectively yielded a total of 12 (4×3) data points for each vocoding channel number, for each participant. As the perceptual effect of noise-vocoding scales is exponential (so for example the increase in intelligibility for normal listeners is much greater between two and four channels than between 20 and 24 channels), we applied a logarithmic (base 2) transformation to the data. The resulting data were then modelled using a Weibull sigmoid, a widely used function for fitting logarithmically scaled data.^45^ Psychometric curves were created for individual participants and a mean curve was created for each diagnostic group using the Matlab psignifit package.^45^ For each function, we report the following parameters: the binaural noise-vocoded speech intelligibility threshold (the number of vocoding channels at which 50% identification of noise-vocoded numbers was achieved); the slope of the function at the threshold point; and lambda (the lapse rate, or number of incorrect responses at maximum performance asymptote),

As the data were not normally distributed, we used nonparametric Kruskal Wallis tests to analyse psychometric parameters. Where the omnibus test was significant, we conducted Dunn’s tests to conduct pairwise comparisons between participant groups. We assessed the relationship of noise-vocoded speech intelligibility threshold to forward digit span over the whole patient cohort, using Spearman’s correlation; here, digit span provides a metric of each patient’s overall ability to repeat (hear, hold in short term memory and articulate) natural spoken numbers. We further used Spearman’s correlation to assess the relationship of intelligibility threshold to general demographic (age, sex), clinical (symptom duration, Mini-Mental State Examination (MMSE) score), executive performance (WASI Matrices) and auditory perceptual (pure tone audiometry, phonemic pairs discrimination on the Psycholinguistic Assessment of Language Processing in Aphasia (PALPA)-3 subtest) measures, over the combined patient cohort.

Finally, receiver operating characteristic (ROC) curves were derived to assess the overall diagnostic utility of noise-vocoded speech comprehension in distinguishing each patient group from healthy controls. The binary classifier used was the 50% speech intelligibility threshold obtained from each psychometric function. The area under the ROC curve (AUC) was calculated for each syndromic group using parametric estimates in the pROC R package.^46,47^

### Brain image acquisition and analysis

Volumetric brain MR images were acquired for 25 patients in a 3 Tesla Siemens Prisma MRI scanner, using a 32-channel phased array head coil and following a T1-weighted sagittal 3D magnetisation prepared rapid gradient echo (MPRAGE) sequence (TE = 2.9 ms, TI = 900 ms, TR = 2200 ms), with dimensions 256 mm x 256 mm x 208 mm and voxel size 1.1 mm x 1.1 mm x 1.1 mm.

For the VBM analysis, patients’ brain images were first pre-processed and normalised to MNI space using SPM12 software (http://www.fil.ion.ucl.ac.uk/spm/software/spm12/) and the DARTEL toolbox with default parameters running under Matlab R2014b. Images were smoothed using a 6-mm full-width at half-maximum Gaussian (FWHM) kernel. To control for individual differences in total (pre-morbid) brain size, total intracranial volume was calculated for each participant by summing white matter, grey matter and cerebrospinal fluid volumes post segmentation.^48^ An explicit brain mask was created using an automatic mask-creation strategy designed previously.^49^ A study-specific mean brain template image upon which to overlay statistical parametric maps was created by warping all patients’ native-space whole-brain images to the final DARTEL template and using the ImCalc function to generate an average of these images.

We assessed grey matter associations of noise-vocoded speech intelligibility threshold over the combined patient cohort. Voxel-wise grey matter intensity was modelled as a function of performance threshold in a multiple regression design, incorporating age, total intracranial volume and diagnostic group membership as covariates. Statistical parametric maps were assessed at peak-level significance threshold p<0.05, after family-wise error (FWE) correction for multiple voxel-wise comparisons within five pre-defined regions of interest, based on prior neuroanatomical hypotheses. These regions comprised left planum temporale,^36,37^ left angular gyrus,^38-40^ left anterior superior temporal gyrus,^38,50,51^ left inferior frontal gyrus ^38,41,50^ and left cingulate gyrus.^38,52^ Anatomical volumes were derived from Oxford-Harvard cortical maps ^53^ and are shown in Figure S2 in Supplementary Material online.

## RESULTS

### General participant group characteristics

Participant groups did not differ significantly in age, sex distribution, handedness or years of formal education (all p>0.05, Table 1). Patient groups did not differ in mean symptom duration (p=0.09) but did differ in MMSE score (H(3)=11.3, p=0.01; see Table 1), the AD group performing worse than the nfvPPA (z=-3.22, p=0.001) and svPPA (z=-2.10, p=0.04) groups. General neuropsychological profiles were in keeping with syndromic diagnosis for each patient group (Table 1). Pure tone audiometry (in the participant subcohort assessed in-person) revealed no substantial peripheral hearing deficits nor any significant differences between participant groups. Basic speech discrimination (assessed using the PALPA-3 phoneme discrimination subtest) performance did not differ significantly from the healthy control group for any of the PPA syndromic groups.

### Experimental behavioural data

Psychometric parameters for the participant groups are presented in Table 2 and noise-vocoded speech intelligibility thresholds for individual participants are plotted in Figure 1. Group mean psychometric functions are presented in Figure 2 and ROC curves for the patient groups versus the healthy control group in Figure 3. Exclusion of two upper bound outliers (>97.5 quantile) in parallel analyses left the results qualitatively unaltered. Results from the full dataset are accordingly reported in-text below; parallel analyses with outliers removed are reported in Supplementary Materials.

**Table 2.**
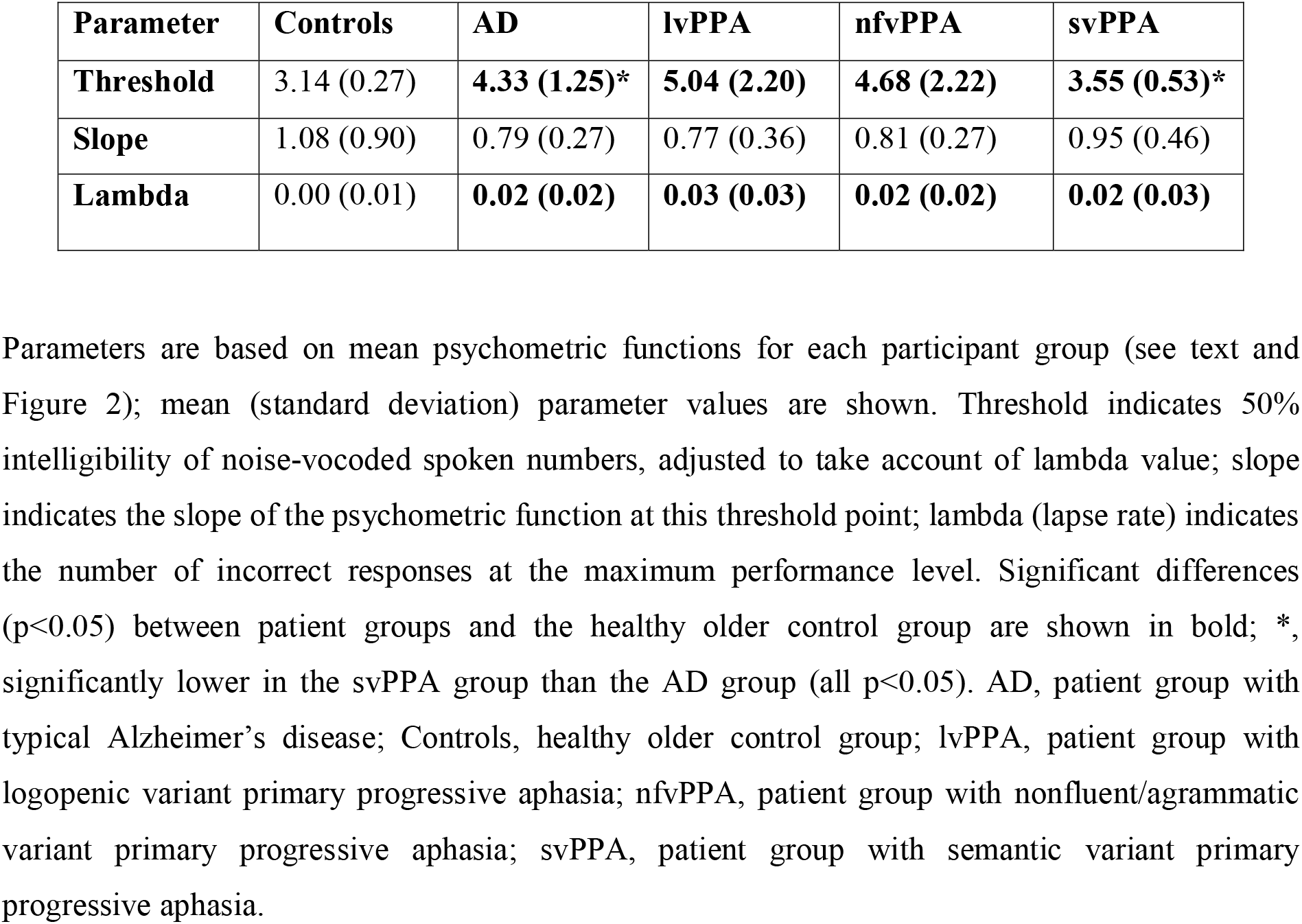
Psychometric function parameters for comprehension of noise-vocoded speech in each participant group

**Figure 1.**
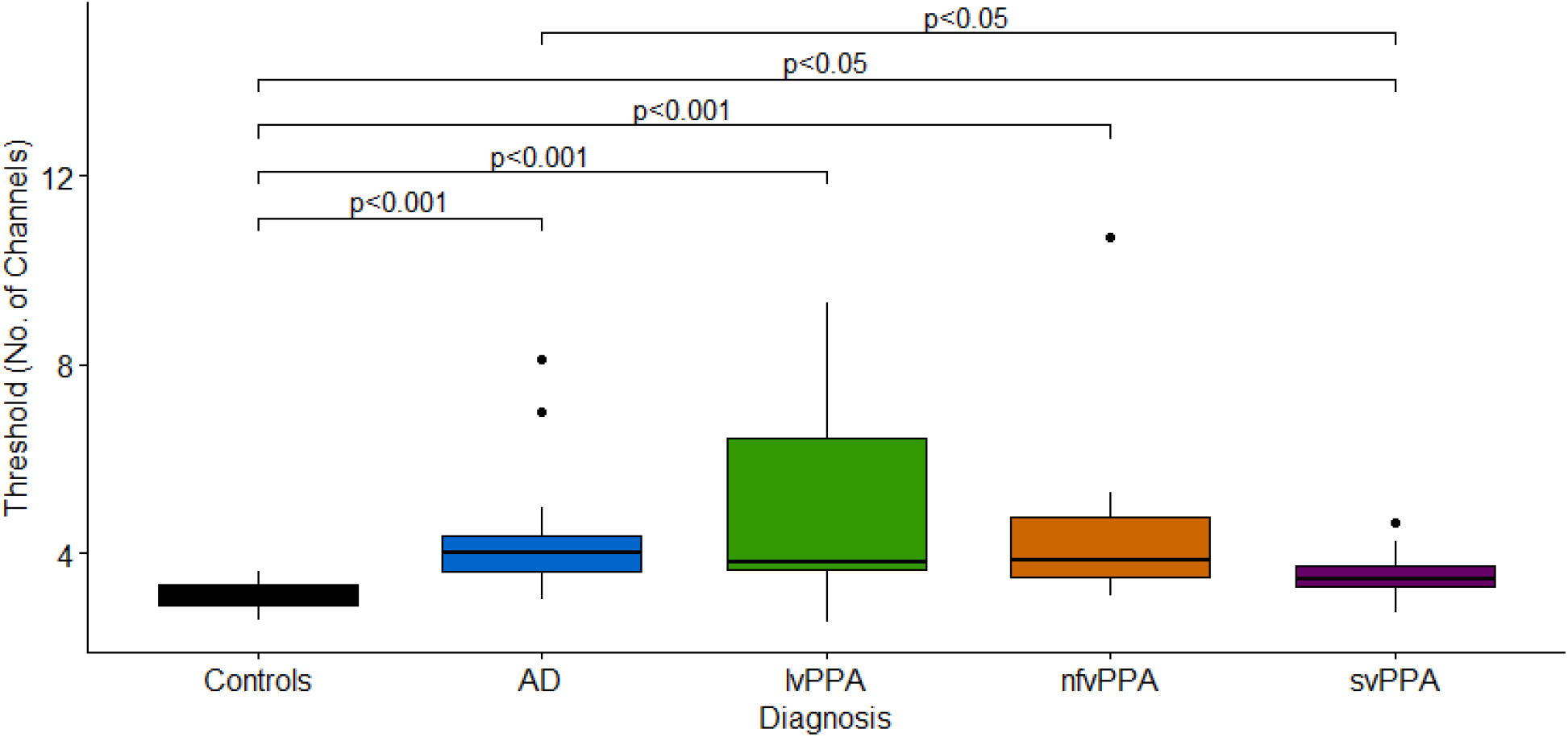
Plots of individual participant thresholds for comprehension of noise-vocoded speech within each diagnostic group. Speech intelligibility threshold values are based on individual psychometric curves for identification of noise-vocoded spoken numbers (see text for details). In this context, threshold corresponds to number of vocoding channels in the speech stimulus at which 50% intelligibility of spoken numbers was achieved, adjusted to take account of lambda value (the upper performance asymptote; see Table 2). The line within each box indicates the median, with the boxes indicating the interquartile interval. AD, patient group with typical Alzheimer’s disease; Control, healthy older control group; lvPPA, patient group with logopenic variant primary progressive aphasia; nfvPPA, patient group with nonfluent variant primary progressive aphasia; svPPA, patient group with semantic variant primary progressive aphasia.

**Figure 2.**
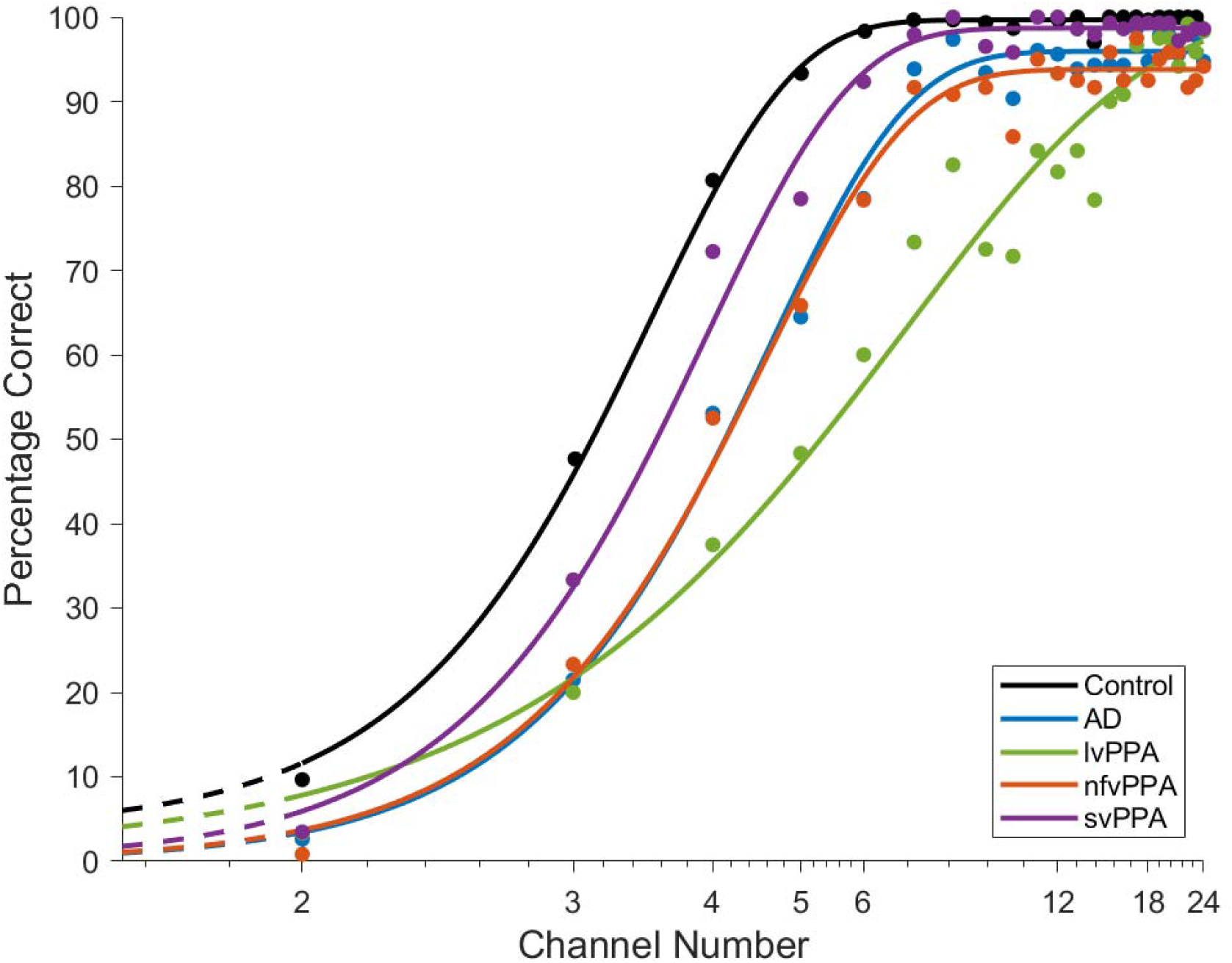
Average psychometric curves for comprehension of noise-vocoded speech in each participant group. The y-axis here shows the percentage of digits identified correctly (from a total of 12 digits) at each noise-vocoding level; the x-axis shows the number of vocoding channels, plotted on a log scale. Mean psychometric functions were created for each diagnostic group (colour coded at lower right; see also text and Table 2); curves have been fitted through values (coloured dots) representing the mean score correct across individual participants in that group at each noise-vocoding level. AD, patient group with typical Alzheimer’s disease; Control, healthy older control group; lvPPA, patient group with logopenic variant primary progressive aphasia; nfvPPA, patient group with nonfluent variant primary progressive aphasia; svPPA, patient group with semantic variant primary progressive aphasia.

**Figure 3.**
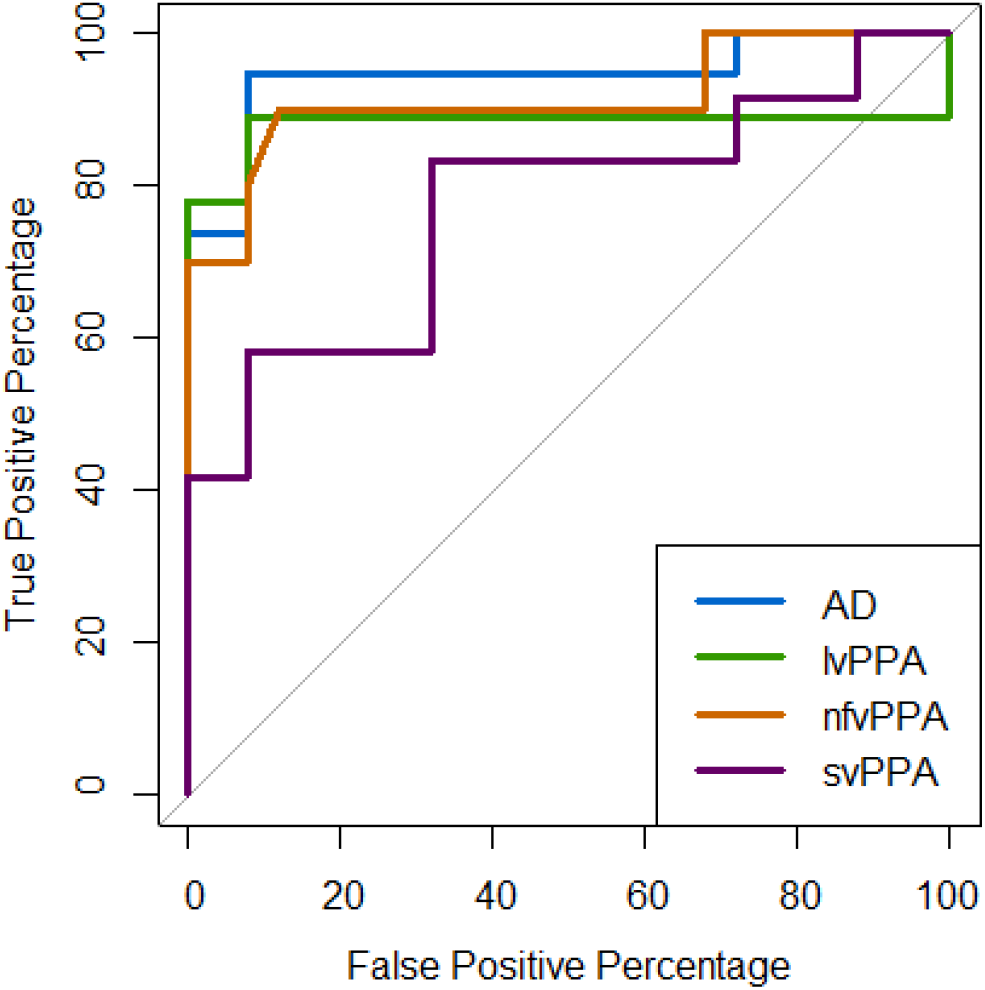
ROC curves for comprehension of noise-vocoded speech in patient groups versus healthy older controls. Receiver operating characteristic (ROC) curves for each syndromic group versus the healthy older control group are shown; the binary classifier used was the speech intelligibility threshold obtained in the psychometric functions (see Table 2 and Figure 2). An area under the curve (AUC) of 1 would correspond to an ideal classifier. AUC values obtained were as follows: Alzheimer’s disease, AUC = 95%; logopenic variant primary progressive aphasia (PPA), AUC = 88%; nonfluent/agrammatic variant PPA, AUC = 91%; semantic variant PPA, AUC = 77%.

There was a significant main effect of diagnostic group on noise-vocoded speech intelligibility threshold (H(4) = 34.35, p<0.001). In post-hoc pairwise group comparisons versus healthy controls, mean intelligibility threshold was significantly elevated in all patient groups: in the lvPPA (z=3.87, p<0.001), nfvPPA (z=3.92, p<0.001), AD (z=5.01, p<0.001) and svPPA (z=2.20, p=0.03) groups. Comparing patient groups, intelligibility threshold was significantly elevated in the AD group compared with the svPPA group (z=2.04, p=0.04). There was no significant effect of diagnostic group on the slope of the psychometric function (p=0.247). There was however a significant main effect of diagnostic group on the lapse rate, lambda (H(4) = 16.75, p=0.002). In post-hoc pairwise group comparisons versus healthy controls, there was a significantly higher lapse rate (more errors made at maximum performance) in all patient groups: in the lvPPA (z=2.95, p=0.003), AD (z=2.61, p=0.009), nfvPPA (z=3.27, p=0.001), and svPPA (z=2.32, p=0.02) groups. There were no significant differences between patient groups for lapse rate.

Individual variability in psychometric parameters within participant groups was substantial (Figure 1, Table 2). Most pertinently, variation in noise-vocoded speech intelligibility threshold was wider in the AD group than in healthy controls and most marked in the lvPPA and nfvPPA groups.

Over the combined patient cohort, noise-vocoded speech intelligibility threshold was not significantly correlated with peripheral hearing function (r=-0.04, p=0.856), phonological discrimination in clear speech (PALPA-3 score; r=-0.25, p=0.185), age (r=0.24, p=0.100) or symptom duration (r=-0.10, p=0.510). Intelligibility threshold in the patient cohort was significantly correlated with WASI Matrices score (r=-0.49, p<0.001), MMSE score (r=-0.53, p<0.001) and forward digit span (r=-0.66, p<0.001). Lapse rate was also significantly correlated with forward digit span across the combined patient cohort (r=-0.34, p=0.017).

Analysis of ROC curves revealed that noise-vocoded speech intelligibility threshold discriminated all patient groups well from healthy controls. Based on AUC values (where a value of 1 would indicate an ideal classifier and values >0.8 a clinically robust discriminator^54,55^), discrimination was ‘excellent’ for the AD group (AUC 0.95) and nfvPPA group (AUC 0.91), ‘good’ for the lvPPA group (AUC 0.88), and ‘fair’ for the svPPA group (AUC 0.77).

### Neuroanatomical data

Statistical parametric maps of grey matter regions associated with speech intelligibility threshold are shown in Figure 4 and local maxima are summarised in Table 3.

**Table 3.** Neuroanatomical associations of noise-vocoded speech intelligibility threshold in the patient cohort

| Region | Cluster size<br>(voxels) | Peak (mm) |  |  | T score | P<sub>FWE</sub> |
| --- | --- | --- | --- | --- | --- | --- |
|  |  | x | y | z |  |  |
| Left planum temporale | 131 | -48 | -31 | 6 | 4.65 | 0.019 |
| Left angular gyrus | 36 | -51 | -61 | 16 | 4.51 | 0.037 |
| Left cingulate gyrus | 142 | -1 | 38 | -5 | 5.68 | 0.012 |

**Figure 4.**
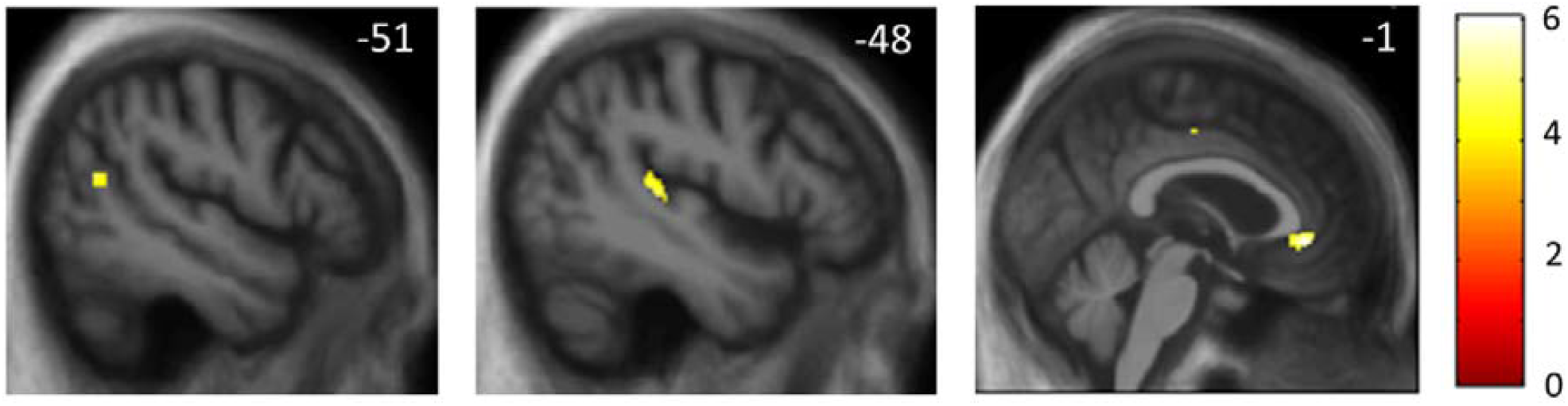
Statistical parametric maps of regional grey matter atrophy associated with elevated noise-vocoded speech intelligibility threshold in the combined patient cohort. Maps are rendered on sagittal sections of the group mean T1-weighted MR image in MRI space, masked using the pre-specified neuroanatomical region of interests (as used in the small volume corrections) and thresholded at p < 0.001 uncorrected for multiple voxel-wise comparisons over the whole brain for display purposes (areas shown were significant at p < 0.05_FWE_ for multiple comparisons within regions of interest). The colour bar (right) codes voxel-wise t-values. All sections are through the left cerebral hemisphere; the plane of each section is indicated using the corresponding MNI coordinate (mm).

Across the combined patient cohort, intelligibility threshold was significantly negatively associated with regional grey matter volume (i.e., associated with grey matter atrophy) in left planum temporale, left angular gyrus, and anterior cingulate gyrus (all p_FWE_ < 0.05 after correction for multiple voxel-wise comparisons within the relevant pre-specified neuroanatomical region of interest).

## DISCUSSION

Here we have shown that perception of acoustically degraded (noise-vocoded) speech is impaired in patients with AD and PPA syndromes relative to healthy older listeners, and further, stratifies syndromes: impairment was most severe in lvPPA and nfvPPA, and significantly more severe in AD than in svPPA. Intelligibility threshold for noise-vocoded speech did not correlate with measures of pure tone detection or phoneme discrimination in clear speech, suggesting that the deficit does not simply reflect a problem with peripheral hearing or elementary speech perception. Individual noise-vocoded speech intelligibility threshold varied widely within the AD, lvPPA and nfvPPA groups. Our findings suggest that elevation in noise-vocoded speech intelligibility threshold in these dementia syndromes captures a central auditory impairment potentially relevant to difficulties in diverse everyday listening situations requiring the decoding of acoustically altered speech signals.

Neuroanatomically, impaired noise-vocoded speech comprehension across dementia syndromes was underpinned by atrophy of left planum temporale, angular gyrus and anterior cingulate gyrus. This cortical network has been shown to be critical for processing speech signals under a range of noisy, daily listening conditions.^5,30,31,40,56^ Planum temporale is likely to play a fundamental role in the deconvolution of complex sound patterns and engagement of neural representations corresponding to phonemes and other auditory objects.^36,37,57^ Angular gyrus mediates the disambiguation of speech signals in challenging listening environments, working memory for speech signals and transcoding of auditory inputs for motor responses, including orienting and repetition.^39,57-60^ Both regions are targeted in AD, lvPPA and nfvPPA ^61-64^ and have been particularly implicated in the pathogenesis of impaired speech perception in these diseases.^27,28,30,65^ The anterior cingulate cortex operates in concert with these more posterior cortical hubs to decode spoken messages under challenging listening conditions,^38,52^ with a more general role in cognitive control and in allocating attentional resources to salient stimuli.^56,66,67^ Reduced activation of the anterior cingulate cortex during tracking of information in degraded speech signals has been demonstrated in nfvPPA and svPPA.^31^

These neuroanatomical considerations suggest that the mechanisms of impaired noise-vocoded speech intelligibility are likely to differ between neurodegenerative syndromes, in keeping with the dissociable processes involved in phoneme recognition.^2^ Noise-vocoding fundamentally reduces the availability of acoustic cues that define phonemes as auditory objects: impaired recognition of these degraded auditory objects could in principle result from deficient encoding of acoustic features, damaged object–level representations (the auditory analogue of ‘apperceptive’ deficits in the visual domain) or impaired top-down, predictive disambiguation based on stored knowledge about speech signal characteristics. In AD and lvPPA, a core deficit of object-level representations has been demonstrated neuropsychologically and electrophysiologically using other procedures that alter acoustic detail in phonemes and nonverbal sounds ^29,31,68,69^; it is therefore plausible that an analogous apperceptive deficit may have impacted the recognition of noise-vocoded phonemes in the AD and lvPPA groups here. In nfvPPA, one previous study of noise-vocoded speech perception has foregrounded the role of inflexible top-down predictive decoding mechanisms instantiated in frontal cortex.^41^ However, this is a clinically, neuroanatomically and neuropathologically diverse syndrome, and involvement of posterior superior temporal cortex engaged in early auditory pattern analysis may constitute a ‘second hit’ to phoneme recognition.^31,68,70,71^ In svPPA, the elevated noise-vocoded intelligibility threshold is *a priori* more likely to reflect reduced activation of semantic mechanisms engaged in the predictive disambiguation of degraded speech signals; and indeed, comprehension of other kinds of acoustically degraded speech signals by patients with svPPA has previously been shown to be sensitive to semantic predictability and to engage anterior cingulate cortex.^27,29,31^

Increasing intelligibility threshold was correlated with digit span over the combined patient cohort. This suggests that verbal working memory limitations may be integrally related to impaired processing of degraded speech, consistent with previous work highlighting the role of working memory in speech perception, particularly in older adults.^72,73^ As working memory demands did not vary across trials and number of vocoding channels, the principal driver of intelligibility threshold is likely to have been the level of acoustic alteration in the speech signal. On the other hand, all patient groups showed an increased lapse rate (i.e., errors unrelated to the stimulus level^45^) at higher vocoding channel numbers (i.e., for minimally noise-vocoded speech signals approaching clear speech). This echoes previous work demonstrating that active listening can be abnormal in lvPPA and nfvPPA even for clear speech and other sounds in quiet.^65,74^ As lapse rate was also correlated with digit span, reduced working memory may well have played a role here, potentially interacting with top-down mechanisms engaged in the predictive processing of speech.^41^ Indeed, frontal processes are likely to play a broader role in the disambiguation of degraded speech signals, including the allocation of attentional and executive resources^75^ and according with the observed correlation here between noise-vocoded speech intelligibility threshold and WASI Matrices score. Taken together, the present findings corroborate the profiles of deficit previously documented in AD and PPA syndromes for comprehension of sinewave speech and phonemic restoration in noise-interrupted speech.^27,29^

Our findings further suggest that markers of noise-vocoded speech comprehension may have diagnostic and biomarker potential. The ROC analysis on the noise-vocoded intelligibility threshold measure (Figure 3) suggests that it would constitute an ‘excellent’ clinical test (corresponding to AUC > 0.9) for discriminating patients with AD and nfvPPA from healthy older individuals.^55^ Additionally, the noise-vocoded intelligibility threshold was correlated with overall disease severity (MMSE score) in the patient cohort. These findings build on a growing body of work suggesting that markers of ‘central’ hearing (auditory cognition) may sensitively signal the functional integrity of cortical regions that are vulnerable to AD and other neurodegenerative pathologies.^5,8,14^ The results of this study could further motivate the development of tailored strategies to help manage hearing difficulties experienced by people with dementia in various daily-life contexts and environments.

This study has limitations that suggest directions for further work. Our noise-vocoding paradigm (based on a step-wise linear progression through channel numbers) was not optimally efficient; an adaptive staircase procedure would reduce testing time and allow individual thresholds to be captured without administering uninformative trials at higher channel numbers. It would be relevant to assess to what extent patients’ comprehension of noise-vocoded speech can be modulated: pharmacologically (in particular, by acetylcholinesterase inhibitors^28^) and/or by perceptual learning, as in healthy listeners.^76-78^ Using another kind of speech degradation (sinewave transformation), we have previously shown that pharmacological and perceptual learning effects may operate in AD and PPA syndromes.^27,28^ To establish how noise-vocoded speech perception and its modulatory factors relate to neural circuit integrity in AD and PPA, functional neuroimaging using techniques such as fMRI and magnetoencephalography will be required to capture dynamic network connectivity engaged by these processes.

From a clinical perspective, this work should be taken forward in several ways. The group sizes here were relatively small: the noise-vocoding paradigm should be extended to larger patient cohorts, which (given the comparative rarity of PPA) will likely entail multi-centre collaboration. Besides corroborating the present group findings, assessment of larger cohorts would allow characterisation of the sources of the wide individual variation within diagnostic groups. There is also a need for prospective, longitudinal studies – both to assess how markers of degraded speech perception relate to disease course and to determine how early such markers may signal underlying neurodegenerative pathology. Auditory measures based on degraded speech comprehension would be well suited to future digital applications and potentially to large-scale screening of populations at risk of incident AD, as well as outcome measures in clinical trials of pharmacotherapies and non-pharmacological interventions.^8,14^ The key next step, however, will be to establish how well measures of degraded speech comprehension correlate with daily-life hearing and communication in AD and other neurodegenerative diseases – using both currently standardised symptom questionnaires and bespoke instruments developed to capture functional hearing disability in dementia. We have previously shown that pure tone audiometry alone is a poor predictor of everyday hearing^79^ while degraded speech performance may have better predictive value in patients with dementia.^80^ There would be considerable clinical value in a quantifiable index of degraded speech perception that could serve as a proxy and predictor of daily life hearing function and disability in major dementias: comprehension of noise-vocoded speech is a promising candidate.

The link between hearing impairment and dementia continues to be debated but presents a major opportunity for earlier diagnosis and intervention. Our findings suggest that the perception of degraded (noise-vocoded) speech captures quantifies central hearing functions beyond sound detection in dementia and stratifies major dementia syndromes. This central hearing index may constitute a proxy for the communication difficulties experienced by patients with AD and PPA under challenging listening conditions in daily life. We hope that this work will motivate further studies to define the diagnostic and therapeutic scope of central hearing measures based on degraded speech perception in these diseases.

## Supporting information

Example Soundfiles

## Data Availability

Data produced in the present study are available upon reasonable request to the authors. The data are not publicly available because they contain information that could compromise the privacy of research participants.

## ACKNOWLEDGEMENTS

We are grateful to all participants for their involvement. We thank Stuart Rosen for helpful advice on the application and analysis of the noise-vocoding paradigm.

## REFERENCES

1. Bregman AS. Auditory scene analysis: The perceptual organization of sound. MIT press; 1994.

2. Goll JC, Crutch SJ, Warren JD. Central auditory disorders: toward a neuropsychology of auditory objects. Current Opinion in Neurology. 2010;23(6):10. doi:10.1097/WCO.0b013e32834027f6

3. Griffiths TD, Warren JD. What is an auditory object? Nature Reviews Neuroscience. 2004;5:5.

4. Lin FR, Metter EJ, O’Brien RJ, Resnick SM, Zonderman AB, Ferrucci L. Hearing Loss and Incident Dementia. Archives of Neurology. 2011;68(2):214–220. doi:10.1001/archneurol.2010.362

5. Jiang J, Benhamou E, Waters S, et al. Processing of Degraded Speech in Brain Disorders. Brain Sciences. 2021;11(394):28. doi:10.3390/brainsci11030394

6. Gates GA, Anderson ML, Feeney MP, McCurry SM, Larson EB. Central Auditory Dysfunction in Older Persons With Memory Impairment or Alzheimer Dementia. Archives of Otolaryngology–Head & Neck Surgery. 2008;134(7):771. doi:10.1001/archotol.134.7.771

7. Gates GA, Anderson ML, McCurry SM, Feeney MP, Larson EB. Central auditory dysfunction as a harbinger of Alzheimer dementia. Arch Otolaryngol Head Neck Surg. Apr 2011;137(4):390–5. doi:10.1001/archoto.2011.28

8. Johnson JCS, Marshall CR, Weil RS, Bamiou D-E, Hardy CJD, Warren JD. Hearing and dementia: from ears to brain. Brain. 2021;144(2):391–401. doi:10.1093/brain/awaa429

9. Livingston G, Sommerlad A, Orgeta V, et al. Dementia prevention, intervention, and care. The Lancet. 2017;390(10113):2673–2734. doi:10.1016/s0140-6736(17)31363-6

10. Griffiths TD, Lad M, Kumar S, et al. How Can Hearing Loss Cause Dementia? Neuron. 2020/11/11/ 2020;108(3):401–412. doi:https://doi.org/10.1016/j.neuron.2020.08.003

11. Powell DS, Oh ES, Reed NS, Lin FR, Deal JA. Hearing Loss and Cognition: What We Know and Where We Need to Go. Front Aging Neurosci. 2021;13:769405. doi:10.3389/fnagi.2021.769405

12. Loughrey DG, Kelly ME, Kelley GA, Brennan S, Lawlor BA. Association of Age-Related Hearing Loss With Cognitive Function, Cognitive Impairment, and Dementia: A Systematic Review and Meta-analysis. JAMA Otolaryngology–Head & Neck Surgery. 2018;144(2):115–126. doi:10.1001/jamaoto.2017.2513

13. Pronk M, Lissenberg-Witte BI, van der Aa HPA, et al. Longitudinal Relationships Between Decline in Speech-in-Noise Recognition Ability and Cognitive Functioning: The Longitudinal Aging Study Amsterdam. J Speech Lang Hear Res. Apr 26 2019;62(4s):1167–1187. doi:10.1044/2018_jslhr-h-ascc7-18-0120

14. Stevenson JS, Clifton L, Kuźma E, Littlejohns TJ. Speech-in-noise hearing impairment is associated with an increased risk of incident dementia in 82,039 UK Biobank participants. Alzheimer’s & Dementia. 2022;18(3):445–456. doi:https://doi.org/10.1002/alz.12416

15. Golden HL, Agustus JL, Goll JC, et al. Functional neuroanatomy of auditory scene analysis in Alzheimer’s disease. Neuroimage Clin. 2015;7:699–708. doi:10.1016/j.nicl.2015.02.019

16. Bouma A, Gootjes L. Effects of attention on dichotic listening in elderly and patients with dementia of the Alzheimer type. Brain Cogn. Jul 2011;76(2):286–93. doi:10.1016/j.bandc.2011.02.008

17. Idrizbegovic E, Hederstierna C, Dahlquist M, Kämpfe Nordström C, Jelic V, Rosenhall U. Central auditory function in early Alzheimer’s disease and in mild cognitive impairment. Age and Ageing. 2011;40(2):249–254. doi:10.1093/ageing/afq168

18. Utoomprurkporn N, Hardy CJD, Stott J, Costafreda SG, Warren J, Bamiou DE. “The Dichotic Digit Test” as an Index Indicator for Hearing Problem in Dementia: Systematic Review and Meta-Analysis. J Am Acad Audiol. Oct 2020;31(9):646–655. doi:10.1055/s-0040-1718700

19. Goll JC, Kim LG, Ridgway GR, et al. Impairments of auditory scene analysis in Alzheimer’s disease. Brain. Jan 2012;135(Pt 1):190–200. doi:10.1093/brain/awr260

20. Golden HL, Agustus JL, Nicholas JM, et al. Functional neuroanatomy of spatial sound processing in Alzheimer’s disease. Neurobiology of Aging. 2016;39:154–164.

21. Golden HL, Nicholas JM, Yong KXX, et al. Auditory spatial processing in Alzheimer’s disease. Brain. 2015;138(1):189–202. doi:10.1093/brain/awu337

22. Hardy CJD, Yong KXX, Goll JC, Crutch SJ, Warren JD. Impairments of auditory scene analysis in posterior cortical atrophy. Brain. Sep 1 2020;143(9):2689–2695. doi:10.1093/brain/awaa221

23. Bidelman GM, Howell M. Functional changes in inter- and intra-hemispheric cortical processing underlying degraded speech perception. Neuroimage. Jan 1 2016;124(Pt A):581–590. doi:10.1016/j.neuroimage.2015.09.020

24. Burda AN, Hageman CF, Brousard KT, Miller AL. Dementia and identification of words and sentences produced by native and nonnative English speakers. Percept Mot Skills. Jun 2004;98(3 Pt 2):1359–62. doi:10.2466/pms.98.3c.1359-1362

25. Hailstone JC, Ridgway GR, Bartlett JW, Goll JC, Crutch SJ, Warren JD. Accent processing in dementia. Neuropsychologia. Jul 2012;50(9):2233–44. doi:10.1016/j.neuropsychologia.2012.05.027

26. Fletcher PD, Downey LE, Agustus JL, et al. Agnosia for accents in primary progressive aphasia. Neuropsychologia. Aug 2013;51(9):1709–15. doi:10.1016/j.neuropsychologia.2013.05.013

27. Hardy CJD, Marshall CR, Bond RL, et al. Retained capacity for perceptual learning of degraded speech in primary progressive aphasia and Alzheimer’s disease. Alzheimers Res Ther. Jul 25 2018;10(1):70. doi:10.1186/s13195-018-0399-2

28. Hardy CJD, Hwang YT, Bond RL, et al. Donepezil enhances understanding of degraded speech in Alzheimer’s disease. Ann Clin Transl Neurol. Nov 2017;4(11):835–840. doi:10.1002/acn3.471

29. Jiang J, Johnson JCS, Requena-Komuro M-C, et al. Phonemic restoration in Alzheimer’s disease and semantic dementia: a preliminary investigation. Brain Communications. 2022;4(3)doi:10.1093/braincomms/fcac118

30. Hardy CJD, Agustus JL, Marshall CR, et al. Behavioural and neuroanatomical correlates of auditory speech analysis in primary progressive aphasias. Alzheimer’s Research & Therapy. 2017;9(1)doi:10.1186/s13195-017-0278-2

31. Hardy CJD, Agustus JL, Marshall CR, et al. Functional neuroanatomy of speech signal decoding in primary progressive aphasias. Neurobiol Aging. Aug 2017;56:190–201. doi:10.1016/j.neurobiolaging.2017.04.026

32. Gates GA, Mills JH. Presbycusis. Lancet. Sep 24-30 2005;366(9491):1111–20. doi:10.1016/s0140-6736(05)67423-5

33. Holmes E, Zeidman P, Friston KJ, Griffiths TD. Difficulties with Speech-in-Noise Perception Related to Fundamental Grouping Processes in Auditory Cortex. Cerebral Cortex. 2020;31(3):1582–1596. doi:10.1093/cercor/bhaa311

34. Shannon RV, Zeng FG, Kamath V, Wygonski J, Ekelid M. Speech Recognition with Primarily Temporal Cues. Science. 1995;270(5234):303–304. doi:10.1126/science.270.5234.303

35. Davis MH, Johnsrude IS. Hearing speech sounds: top-down influences on the interface between audition and speech perception. Hear Res. Jul 2007;229(1-2):132–47. doi:10.1016/j.heares.2007.01.014

36. Warren JD, Scott SK, Price CJ, Griffiths TD. Human brain mechanisms for the early analysis of voices. NeuroImage. 2006/07/01/ 2006;31(3):1389–1397. doi:https://doi.org/10.1016/j.neuroimage.2006.01.034

37. Griffiths TD, Warren JD. The planum temporale as a computational hub. Trends in Neurosciences. 2002/07/01/ 2002;25(7):348–353. doi:https://doi.org/10.1016/S0166-2236(02)02191-4

38. Obleser J, Wise RJS, Alex Dresner M, Scott SK. Functional Integration across Brain Regions Improves Speech Perception under Adverse Listening Conditions. The Journal of Neuroscience. 2007;27(9):2283–2289. doi:10.1523/jneurosci.4663-06.2007

39. Hartwigsen G, Golombek T, Obleser J. Repetitive transcranial magnetic stimulation over left angular gyrus modulates the predictability gain in degraded speech comprehension. Cortex. Jul 2015;68:100–10. doi:10.1016/j.cortex.2014.08.027

40. Davis MH, Johnsrude IS. Hierarchical Processing in Spoken Language Comprehension. The Journal of Neuroscience. 2003;23(8):3423–3431. doi:10.1523/jneurosci.23-08-03423.2003

41. Cope TE, Sohoglu E, Sedley W, et al. Evidence for causal top-down frontal contributions to predictive processes in speech perception. Nat Commun. Dec 18 2017;8(1):2154. doi:10.1038/s41467-017-01958-7

42. Dubois B, Feldman HH, Jacova C, et al. Advancing research diagnostic criteria for Alzheimer’s disease: the IWG-2 criteria. The Lancet Neurology. 2014;13(6):614–629. doi:10.1016/s1474-4422(14)70090-0

43. Gorno-Tempini ML, Hillis AE, Weintraub S, et al. Classification of primary progressive aphasia and its variants. Neurology. 2011;76(11):1006–1014. doi:10.1212/wnl.0b013e31821103e6

44. Requena-Komuro M-C, Jiang J, Dobson L, et al. Remote versus face-to-face neuropsychological testing for dementia research: a comparative study in patients with Alzheimer’s disease, patients with frontotemporal dementia, and healthy older individuals. BMJ Open. 2022;

45. Schütt HH, Harmeling S, Macke JH, Wichmann FA. Painfree and accurate Bayesian estimation of psychometric functions for (potentially) overdispersed data. Vision Research. 2016/05/01/ 2016;122:105–123. doi:https://doi.org/10.1016/j.visres.2016.02.002

46. Robin X, Turck N, Hainard A, et al. pROC: an open-source package for R and S+ to analyze and compare ROC curves. BMC Bioinformatics. 2011;12:77. doi:10.1186/1471-2105-12-77

47. Hajian-Tilaki KO, Hanley JA, Joseph L, Collet J-P. A Comparison of Parametric and Nonparametric Approaches to ROC Analysis of Quantitative Diagnostic Tests. Medical Decision Making. 1997;17(1):94–102. doi:10.1177/0272989x9701700111

48. Malone IB, Leung KK, Clegg S, et al. Accurate automatic estimation of total intracranial volume: a nuisance variable with less nuisance. Neuroimage. Jan 1 2015;104:366–72. doi:10.1016/j.neuroimage.2014.09.034

49. Ridgway GR, Omar R, Ourselin S, Hill DLG, Warren JD, Fox NC. Issues with threshold masking in voxel-based morphometry of atrophied brains. NeuroImage. 2009/01/01/ 2009;44(1):99–111. doi:https://doi.org/10.1016/j.neuroimage.2008.08.045

50. Hervais-Adelman AG, Carlyon RP, Johnsrude IS, Davis MH. Brain regions recruited for the effortful comprehension of noise-vocoded words. Language and Cognitive Processes. 2012;27(7-8):1145–1166. doi:10.1080/01690965.2012.662280

51. Scott SK, Rosen S, Lang H, Wise RJS. Neural correlates of intelligibility in speech investigated with noise vocoded speech—A positron emission tomography study. The Journal of the Acoustical Society of America. 2006;120(2):1075–1083. doi:10.1121/1.2216725

52. Gennari SP, Millman RE, Hymers M, Mattys SL. Anterior paracingulate and cingulate cortex mediates the effects of cognitive load on speech sound discrimination. NeuroImage. 2018/09/01/ 2018;178:735–743. doi:https://doi.org/10.1016/j.neuroimage.2018.06.035

53. Desikan RS, Ségonne F, Fischl B, et al. An automated labeling system for subdividing the human cerebral cortex on MRI scans into gyral based regions of interest. NeuroImage. 2006/07/01/ 2006;31(3):968–980. doi:https://doi.org/10.1016/j.neuroimage.2006.01.021

54. Ohman EM, Granger CB, Harrington RA, Lee KL. Risk Stratification and Therapeutic Decision Making in Acute Coronary Syndromes. JAMA. 2000;284(7):876–878. doi:10.1001/jama.284.7.876

55. Carter JV, Pan J, Rai SN, Galandiuk S. ROC-ing along: Evaluation and interpretation of receiver operating characteristic curves. Surgery. 2016/06/01/ 2016;159(6):1638–1645. doi:https://doi.org/10.1016/j.surg.2015.12.029

56. Wild CJ, Yusuf A, Wilson DE, Peelle JE, Davis MH, Johnsrude IS. Effortful Listening: The Processing of Degraded Speech Depends Critically on Attention. Journal of Neuroscience. 2012;32(40):14010–14021. doi:10.1523/jneurosci.1528-12.2012

57. Warren JE, Wise RJ, Warren JD. Sounds do-able: auditory-motor transformations and the posterior temporal plane. Trends Neurosci. Dec 2005;28(12):636–43. doi:10.1016/j.tins.2005.09.010

58. Shahin AJ, Bishop CW, Miller LM. Neural mechanisms for illusory filling-in of degraded speech. NeuroImage. 2009;44(3):1133–1143. doi:10.1016/j.neuroimage.2008.09.045

59. Obleser J, Kotz SA. Expectancy constraints in degraded speech modulate the language comprehension network. Cereb Cortex. Mar 2010;20(3):633–40. doi:10.1093/cercor/bhp128

60. Golestani N, Hervais-Adelman A, Obleser J, Scott SK. Semantic versus perceptual interactions in neural processing of speech-in-noise. NeuroImage. 2013/10/01/ 2013;79:52–61. doi:https://doi.org/10.1016/j.neuroimage.2013.04.049

61. Lombardi J, Mayer B, Semler E, et al. Quantifying progression in primary progressive aphasia with structural neuroimaging. Alzheimer’s & Dementia. 2021;17(10):1595–1609. doi:10.1002/alz.12323

62. Ruksenaite J, Volkmer A, Jiang J, et al. Primary Progressive Aphasia: Toward a Pathophysiological Synthesis. Current Neurology and Neuroscience Reports. 2021;21(3)doi:10.1007/s11910-021-01097-z

63. Bejanin A, Schonhaut DR, La Joie R, et al. Tau pathology and neurodegeneration contribute to cognitive impairment in Alzheimer’s disease. Brain. 2017;140(12):3286–3300. doi:10.1093/brain/awx243

64. Giannini LAA, Irwin DJ, McMillan CT, et al. Clinical marker for Alzheimer disease pathology in logopenic primary progressive aphasia. Neurology. 2017;88(24):2276–2284. doi:10.1212/wnl.0000000000004034

65. Johnson JCS, Jiang J, Bond RL, et al. Impaired phonemic discrimination in logopenic variant primary progressive aphasia. Ann Clin Transl Neurol. Jul 2020;7(7):1252–1257. doi:10.1002/acn3.51101

66. Shenhav A, Botvinick Matthew M, Cohen Jonathan D. The Expected Value of Control: An Integrative Theory of Anterior Cingulate Cortex Function. Neuron. 2013/07/24/ 2013;79(2):217–240. doi:https://doi.org/10.1016/j.neuron.2013.07.007

67. Abutalebi J, Della Rosa PA, Green DW, et al. Bilingualism Tunes the Anterior Cingulate Cortex for Conflict Monitoring. Cerebral Cortex. 2011;22(9):2076–2086. doi:10.1093/cercor/bhr287

68. Goll JC, Kim LG, Hailstone JC, et al. Auditory object cognition in dementia. Neuropsychologia. Jul 2011;49(9):2755–65. doi:10.1016/j.neuropsychologia.2011.06.004

69. Stalpaert J, Miatton M, Sieben A, Langenhove TV, Mierlo Pv, Letter MD. The Electrophysiological Correlates of Phoneme Perception in Primary Progressive Aphasia: A Preliminary Case Series. Frontiers in Human Neuroscience. 2021;doi:https://doi.org/10.3389/fnhum.2021.618549

70. Goll JC, Crutch SJ, Loo JH, et al. Non-verbal sound processing in the primary progressive aphasias. Brain. Jan 2010;133(Pt 1):272–85. doi:10.1093/brain/awp235

71. Grube M, Bruffaerts R, Schaeverbeke J, et al. Core auditory processing deficits in primary progressive aphasia. Brain. Jun 2016;139(Pt 6):1817–29. doi:10.1093/brain/aww067

72. Meister H, Schreitmüller S, Grugel L, Beutner D, Walger M, Meister I. Examining Speech Perception in Noise and Cognitive Functions in the Elderly. American Journal of Audiology. 2013;22(2):310–312. doi:doi:10.1044/1059-0889(2012/12-0067)

73. Millman RE, Mattys SL. Auditory Verbal Working Memory as a Predictor of Speech Perception in Modulated Maskers in Listeners With Normal Hearing. Journal of Speech, Language, and Hearing Research. 2017;60(5):1236–1245. doi:doi:10.1044/2017_JSLHR-S-16-0105

74. Hardy CJD, Frost C, Sivasathiaseelan H, et al. Findings of Impaired Hearing in Patients With Nonfluent/Agrammatic Variant Primary Progressive Aphasia. JAMA Neurol. May 1 2019;76(5):607–611. doi:10.1001/jamaneurol.2018.4799

75. Peelle JE. Listening Effort: How the Cognitive Consequences of Acoustic Challenge Are Reflected in Brain and Behavior. Ear Hear. Mar/Apr 2018;39(2):204–214. doi:10.1097/aud.0000000000000494

76. Davis MH, Johnsrude IS, Hervais-Adelman A, Taylor K, McGettigan C. Lexical information drives perceptual learning of distorted speech: evidence from the comprehension of noise-vocoded sentences. J Exp Psychol Gen. May 2005;134(2):222–41. doi:10.1037/0096-3445.134.2.222

77. Sohoglu E, Davis MH. Perceptual learning of degraded speech by minimizing prediction error. Proc Natl Acad Sci U S A. Mar 22 2016;113(12):E1747–56. doi:10.1073/pnas.1523266113

78. Hervais-Adelman A, Davis MH, Johnsrude IS, Carlyon RP. Perceptual learning of noise vocoded words: Effects of feedback and lexicality. Journal of Experimental Psychology: Human Perception and Performance. 2008;34(2):460–474. doi:10.1037/0096-1523.34.2.460

79. Utoomprurkporn N, Stott J, Costafreda SG, Bamiou DE. Lack of Association between Audiogram and Hearing Disability Measures in Mild Cognitive Impairment and Dementia: What Audiogram Does Not Tell You. Healthcare (Basel). Jun 20 2021;9(6)doi:10.3390/healthcare9060769

80. Johnson JCS. Hearing impairment in dementia: defining deficits and assessing impact. UCL; 2021.

81. Audiology BSo. Recommended Procedure: Pure-tone air-conduction and bone-conduction threshold audiometry with and without masking. 2018.

82. Finger H, Goeke C, Diekamp D, Standvoß K, König P. LabVanced: a unified JavaScript framework for online studies. 2017:

83. Bench J, Kowal Å, Bamford J. The Bkb (Bamford-Kowal-Bench) Sentence Lists for Partially-Hearing Children. British Journal of Audiology. 1979/01/01 1979;13(3):108–112. doi:10.3109/03005367909078884

