## Supplementary Materials for "Comprehension of acoustically degraded speech in Alzheimer’s disease and primary progressive aphasia"

**Assessment of peripheral hearing function**

A standard clinical audiometer protocol was used to assess participants’ peripheral hearing of pure tones at set frequencies ^81^. Using a GSI Audiostar Pro^TM^ audiometer (<https://www.guymark.com/product-information/hearing-assessment/audiometers/clinical/gsi-audiostar-pro>) and calibrated GSI Audiostar Pro headphones with noise-reducing ear-cups, steady tones of 250, 500, 1000, 2000, 4000 and 8000Hz were presented separately to each of the participant’s ears. At each frequency, the participant indicated (verbally or by gesture) when they first heard a noise. The decibel hearing level was set typically at 50 dB, with decreases of 10 dB if they could hear the tone, and increases of 5dB if they could not.. For each participant, a composite peripheral hearing score was created by calculating the mean threshold across all frequencies in the best ear.

**Synthesis of noise-vocoded stimuli**

Raw speech recordings were noise-vocoded using a script written by Chris Darwin: <http://www.lifesci.sussex.ac.uk/home/Chris_Darwin/Praatscripts/Shannon>. The logspace function in Matlab was used to calculate log spacing between 50 and 8000Hz, corresponded to logarithmically-spaced frequency bands that served as the basis for the noise-vocoding algorithm. The algorithm was run iteratively to generate speech stimuli with between 1 and 24 frequency bands (‘channels’). Average (rms) stimulus intensity was constant for all stimuli, as fixed in Matlab. All stimuli were windowed with 20ms onset-offset temporal ramps to prevent click artefacts.

**Remote testing procedure**

Thirty participants (four healthy controls, nine patients with AD, seven with lvPPA, six with nfvPPA and four with svPPA) were assessed remotely via a video link, due to the COVID-19 pandemic. We have detailed the design and implementation of our remote neuropsychological assessment protocol elsewhere ^44^.

An initial session was conducted via Zoom to accustom participants to the remote testing format, check the screen and sound sharing options on Zoom, and that the quality of the participant’s internet connection was acceptable. Participants were permitted to use their preferred technology interface (computers/tablets; smartphones were not allowed, to ensure good screen visibility). Remote assessments were scheduled to ensure testing could be completed in a quiet environment with minimal distractions, and the device volume was set to a comfortable level by each participant or their caregiver. The experiment was implemented onto Labvanced for administration.^82^ Most participants listened in free field, over their device speakers (only a few used headphones); of the seven healthy control participants who performed the experiment both in person and remotely, only one used headphones for the remote session.

A further audibility check was used for each participant through the Bamford-Kowal-Bench ^83^ list. The spoken BKB sentences were delivered online using Labvanced ^82^; a perfect score of the final 3 items on the test was required to go on to the remote testing session proper (this allowed each participant and/or caregiver to manually adjust the volume to a comfortable level for clear audibility). Most participants (97%) performed at ceiling; none was rejected based on their BKB performance.

Each patient’s caregiver was required to be available during each research session in case of any problems using the equipment. In practice, however, no major technological issues arose during the remote testing sessions.

The remote experimental procedure was otherwise identical to the in-person procedure.

Performance profiles in seven healthy control participants who performed the experiment both in person (via headphones) and remotely (in free-field) were very similar (see Figure S3), justifying combining participants tested in person and remotely in the main analysis.

**Experimental behavioural data results: parallel analyses after outlier removal**

There was a significant main effect of diagnostic group on noise-vocoded speech intelligibility threshold (H(4) = 32.89, p<0.001). In post-hoc pairwise group comparisons versus healthy controls, intelligibility threshold was significantly elevated in all patient groups, most markedly in the lvPPA group (z=3.48, p<0.001), followed by the nfvPPA (z=3.55, p<0.001), AD (z=5.15, p<0.001) and svPPA (z=2.26, p=0.02) groups. Comparing patient groups, intelligibility threshold was significantly elevated in the AD group (z=2.10, p=0.04) compared with the svPPA group. There was no significant effect of diagnostic group on the slope of the psychometric function (p=0.320). There was, however, a significant main effect of diagnostic group on the lapse rate, lambda (H(4) = 19.77, p=0.001). In post-hoc pairwise group comparisons versus healthy controls, there was a significantly higher lapse rate (more errors made at maximum performance) in all patient groups: this elevation was most marked in the lvPPA group (z=3.26, p=0.001) and somewhat less marked (and comparable) in the AD (z=2.60, p=0.009), nfvPPA (z=3.56, p<0.001), and svPPA (z=2.31, p=0.02) groups. No significant difference was seen between patient groups on lapse rate.

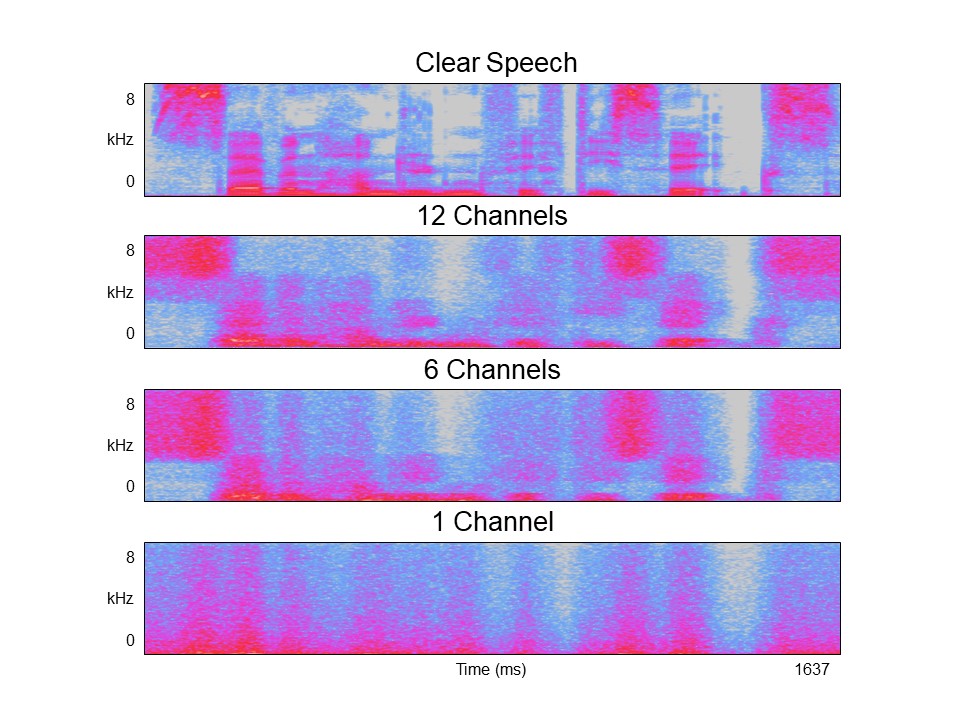

**Figure S1.** Spectrograms of clear (natural) speech and noise-vocoded speech resampled with differing numbers of discrete frequency bands (12 channels, 6 channels and 1 channel). The verbal message in each case is the spoken number ‘seven hundred and fifty-six’. The spectrogram provides a visual indication of how the energy in different frequency bands of the speech signal (plotted on y axis) changes over time (plotted on x-axis). Reducing the number of channels (frequency bands) reduces progressively the amount of spectrotemporal fine structure in the speech signal. Example stimuli are available in Supplementary Materials online.

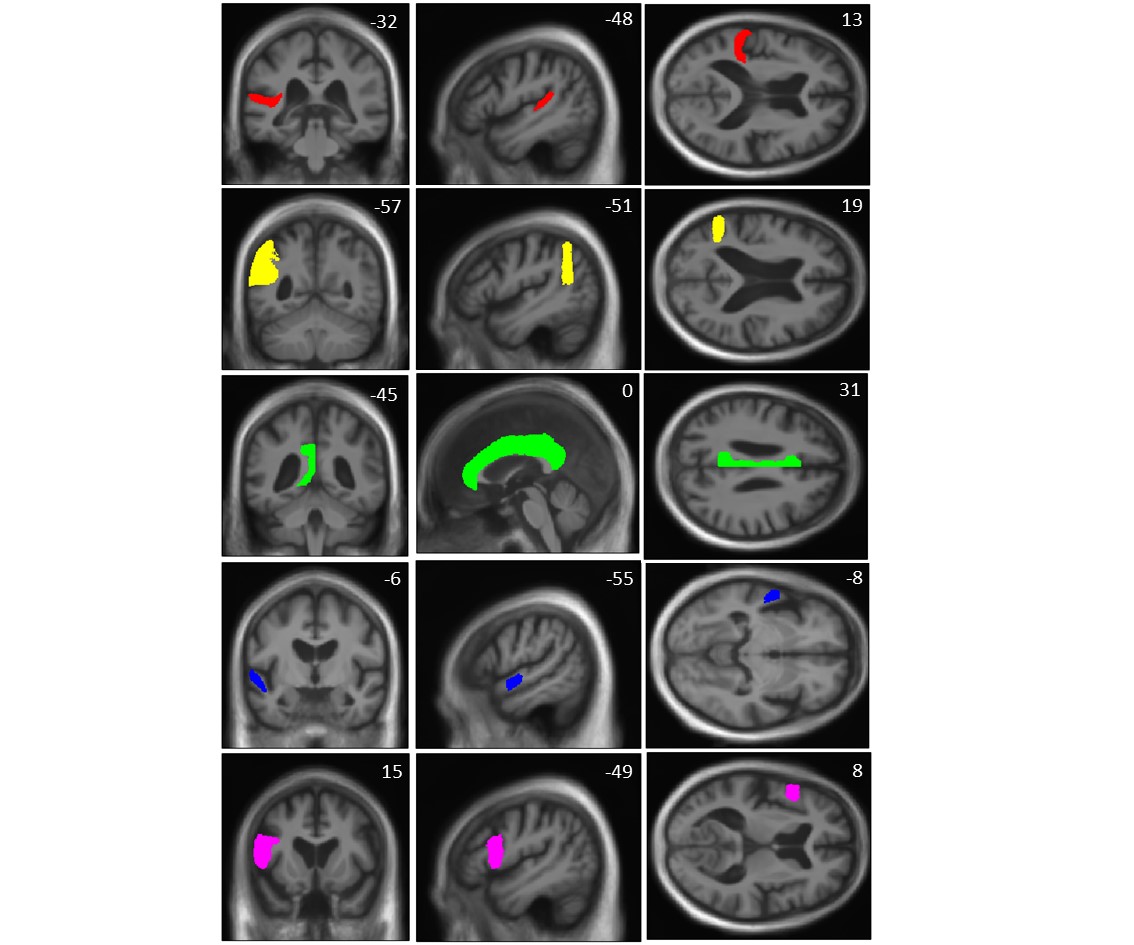

**Figure S2.** Representative sections of neuroanatomical regions in the left cerebral hemisphere that were used for multiple voxel-wise comparisons correction in region-of-interest analyses (see text). Regions are rendered on coronal (left), sagittal (middle) and axial (right) sections of the mean normalised brain template for the patient cohort; MNI coordinates of the plane of each section are shown. The neuroanatomical regions comprise left planum temporale (red), left angular gyrus (yellow), left cingulate gyrus (green), left anterior superior temporal gyrus (blue), and left inferior frontal gyrus (purple).

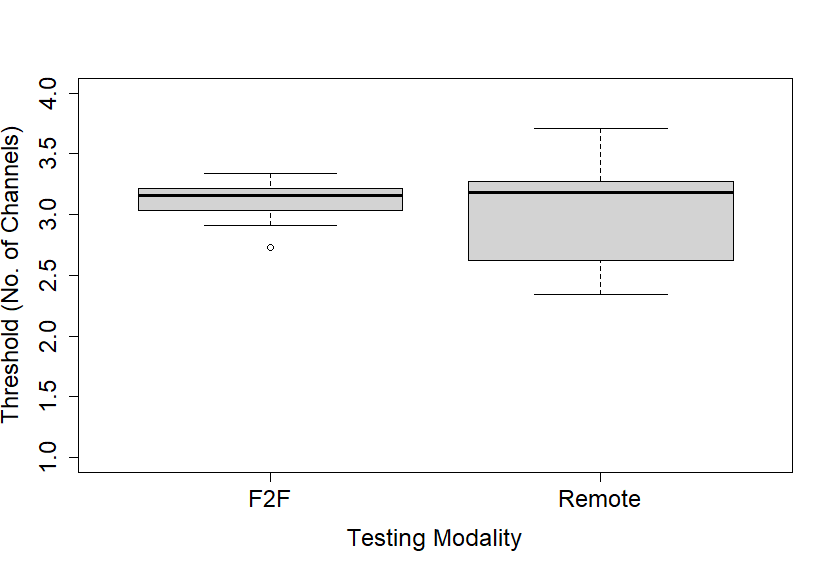

**Figure S3.** Comparison of healthy older controls’ performance on comprehension of noise-vocoded speech for in-person vs remote testing sessions. Seven healthy older control participants performed the noise-vocoded spoken number identification task both face-to-face (F2F) at the research centre and remotely in their home environments, approximately 20 months later. Box plots show the mean and standard deviation (whiskers indicate the full range) of speech intelligibility thresholds in each session. Threshold differences between the two sessions were non-significant (F2F mean = 3.105; Remote mean = 3.003; p=0.69).
